# Patient needs and expectations regarding cognitive impairments in long COVID: perspectives of young and older adults in the UK

**DOI:** 10.64898/2025.12.17.25342465

**Authors:** UK Long COVID Cognitive Experience Research Group, Dan Shan, Trevor J. Crawford, Carol Holland

## Abstract

**Objective:** To explore how younger and older adults in the UK experience, manage, and seek treatment for long COVID-related cognitive impairments, and to identify their treatment preferences and expectations.

**Design:** Qualitative study using Charmaz’s (2014) constructivist grounded theory methodology.

**Setting:** Online, semi-structured interviews conducted with UK adults reporting long COVID-related cognitive symptoms, recruited via patient support groups and research panels.

**Participants:** Twenty-one adults with long COVID cognitive impairments: 10 younger participants aged 25–38 years and 11 older participants aged 60–72 years.

**Main outcome measures:** Patients’ lived experiences, coping strategies, healthcare interactions, and treatment preferences and expectations for non-pharmacological and pharmacological interventions, analysed through iterative coding, constant comparison, and theory generation.

**Results:** In the absence of effective treatments, both age groups relied heavily on self-management strategies (e.g., memory aids, structured routines, pacing). Interactions with healthcare were characterised by validation from some clinicians but widespread frustration at the lack of treatment options. Younger adults were more proactive in seeking experimental therapies and clinical trials, while older adults emphasised pragmatic adaptation, independence, and cautious optimism. Across groups, participants preferred non-pharmacological interventions (e.g., cognitive rehabilitation) but also expressed hope for biomedical treatments. The central process identified was “striving for agency” in the face of ongoing cognitive difficulties.

**Conclusions:** This study highlights the urgent unmet need for evidence-based interventions to address long COVID-related cognitive impairment. Health services should provide practical cognitive rehabilitation and support, and clinicians should acknowledge and validate patients’ cognitive struggles rather than dismissing them as normal ageing or purely psychogenic in origin. Research into therapeutics (e.g. cognitive training programs, pharmacotherapies) is urgently desired by the participants in this study. Effective solutions will need to be holistic and individualised – addressing not only memory and concentration deficits but also the psychological and social challenges associated with long COVID cognitive impairment in different age groups.

**WHAT IS ALREADY KNOWN ON THIS TOPIC:**

- A large proportion of people with long COVID experience persistent cognitive difficulties, often described as “brain fog,” with substantial impact on daily functioning and employment.
- Most studies have focused on symptom prevalence, biological mechanisms, or broad psychosocial consequences.
- Little is known about patients’ treatment preferences and expectations, and almost no research has explored generational differences between younger and older adults.

**WHAT THIS STUDY ADDS:**

- To our knowledge, this is the first qualitative study in the UK to compare younger and older adults’ perspectives on managing long COVID-related cognitive impairment.
- Both groups described “striving for agency” through self-management, validation-seeking in healthcare, and balancing preferences for pharmacological and non-pharmacological treatments.
- Younger adults were more proactive in seeking experimental therapies and trial participation, while older adults emphasised pragmatic adaptation and maintaining independence.

**HOW THIS STUDY MIGHT AFFECT RESEARCH, PRACTICE, OR POLICY:**

- Healthcare services should expand access to cognitive rehabilitation and psychosocial support, while clinicians should validate patients’ experiences rather than attributing them to ageing or stress.
- Future research must test both pharmacological and non-pharmacological interventions, with study designs informed by patient priorities and generational needs.
- Policymakers and service providers should tailor care pathways to life stage: supporting younger adults with workplace accommodations and research opportunities, while providing older adults with reassurance, independence-focused care, and monitoring for possible accelerated cognitive ageing.

## 1 Introduction

Prior evidence suggests that a substantial proportion of individuals with long COVID (post-acute sequelae of SARS-CoV-2 infection, or PASC) can experience persistent difficulties with memory, attention, and executive function, often described as “brain fog”, for months following the acute phase of infection, with cognitive symptoms reported as among the most frequent and debilitating, second only to fatigue.^1^ For example, an international survey of 3,762 long COVID patients found that 88% reported cognitive dysfunction, with concentration and memory problems often persisting beyond six months (in many cases significantly impairing work capacity).^2^ Neuroimaging evidence further demonstrates structural and functional correlates (e.g. loss of grey matter in fronto-limbic networks) that may underlie the “brain fog” phenomenon.^3^ These deficits could drastically undermine an individual’s ability to work, study, and carry out daily activities; one survey found that 45% of long COVID patients had to reduce their work hours and 22% were still not working at all several months after their initial COVID-19 infection.^2^ Long COVID cognitive impairment therefore poses not only a medical concern but also a socioeconomic challenge, contributing substantially to disability and lost productivity, especially in areas already facing disproportionate health burdens and socioeconomic inequalities.^4–6^

Long COVID’s cognitive effects have been documented across a wide range of populations, but there has been little attention to potential differences in how younger versus older adults experience and manage these symptoms. On one hand, older adults may be more vulnerable to severe neurologic complications of COVID-19, partly attributable to potential comorbidities and age-related physiological changes, while their post-COVID cognitive complaints are at risk of being misattributed to ‘normal ageing’.^7^ Indeed, many seniors have had post-COVID cognitive issues dismissed as age-related changes.^8,9^ On the other hand, younger adults with long COVID “brain fog” might encounter scepticism of a different kind – employers or peers might think they are too young to have memory problems, and these individuals often feel guilt or anxiety about failing to meet work and family obligations during their prime years.^10^ A prior study of integrative support group visits for long COVID noted that older participants worried their cognitive symptoms would be attributed to old age, whereas younger participants struggled with loss of independence and career setbacks due to “brain fog”.^11^ Despite these contextual differences, both age groups reported remarkably similar symptom profiles (e.g., short-term memory lapses, concentration difficulties, and mental fatigue), and both identified cognitive impairment as a key contributor to reduced quality of life.^12^ Although prior research has compared younger and older adults with long COVID, most have focused on describing symptom prevalence or broad psychosocial themes rather than systematically examining how age groups differ in managing or adapting to cognitive challenges.^8–11^ As a result, what remains underexplored is whether their needs, preferences, and coping strategies for these cognitive issues differ in meaningful ways. Understanding any such differences is important for tailoring individualised rehabilitation approaches – for instance, an older retired person and a young working parent might have different expectations and constraints when seeking treatment for “brain fog”.^6^

Currently, no formally approved pharmacological treatment exists for long COVID-related cognitive impairment.^13^ A multitude of hypotheses for pathophysiology – including neuroinflammation, microvascular coagulation, viral persistence in the central nervous system, and autoimmune processes – have been proposed, and early-stage trials of various interventions are underway.^14^ For example, small case series have reported potential improvements in cognition with repurposed drugs like guanfacine (an α-agonist used for Attention Deficit Hyperactivity Disorder, or ADHD) when combined with N-acetylcysteine,^15^ or with modalities like hyperbaric oxygen therapy,^16^ but robust evidence from larger randomised trials is still lacking. Although drug therapies have not yet demonstrated clear efficacy for post-COVID cognitive symptoms in published trials,^13^ a recent living systematic review concluded that rehabilitative interventions showed the most promise so far: moderate-certainty evidence indicated that structured cognitive-behavioural therapy (CBT) programs could improve concentration and fatigue, and supervised multidisciplinary rehabilitation (combining physical exercise and mental health support) was associated with overall functional benefits in long COVID patients.^13^

Accordingly, clinical guidelines (e.g. the United Kingdom’s National Institute for Health and Care Excellence [NICE] and the United States Centres for Disease Control and Prevention [CDC]) currently emphasise supportive care and self-management strategies – pacing of activities, use of memory aids, stress reduction, and treatment of contributory factors like sleep disturbance or mood disorders – as the mainstay of managing “brain fog” pending further research.^17^ Specialist “long COVID clinics” established in the UK and elsewhere tended to provide multidisciplinary assessment and general advice rather than disease-specific therapies.^18^ Many patients reported difficulty accessing even these rehabilitative services due to high demand or lack of awareness, leaving them to navigate symptoms largely on their own.^19^ Surveys of long COVID populations have highlighted a frequent sentiment that their healthcare needs are not being met and that they resort to crowdsourced tips and trial-and-error approaches to cope with persistent symptoms.^17^

Although quantitative research is delineating the scope of long COVID cognitive deficits, qualitative research can provide critical insight into how patients themselves experience these deficits, how they respond, and what expectations they hold for care and recovery, and such insights are crucial for developing and implementing treatments that patients find acceptable and effective. A few qualitative studies have begun to document the lived experience of long COVID “brain fog” – for instance, a UK focus group study reported that patients felt unable to cope with simultaneous inputs and consequently adopted pragmatic adaptations, including avoiding multitasking and using written checklists.^20^ In-depth interviews conducted by Kingstone et al. revealed that individuals with persistent long COVID symptoms often felt mentally invalidated (i.e., not taken seriously) and experienced a loss of identity due to cognitive and energy limitations, yet they actively sought ways to regain control over their lives.^21^ However, gaps remain in our understanding of patients’ treatment preferences and expectations regarding these cognitive problems. What kinds of support or interventions do long COVID patients believe would help their “foggy” brains? How do they prioritise among pharmacological, technological, or behavioural approaches? How do they envision recovery, and do these expectations diverge between a young adult hoping to return to a cognitively demanding job versus an older adult fearing dementia? These questions have practical significance for designing patient-centred care for long COVID: aligning interventions with what patients are willing to try (or are sceptical of) can improve uptake and outcomes.

In this study, we used a constructivist grounded theory approach to explore deeply how younger and older adults in the UK perceive and manage their long COVID-related cognitive impairments, with a focus on their treatment-related preferences, behaviours, and hopes. Grounded theory is well-suited to generate a conceptual framework from patient experiences, especially for a novel condition like long COVID where predefined models are lacking.^22^ By comparing younger and older participants, we aimed to illuminate any age-related influences on coping strategies or help-seeking. Our goal was to construct a theory that could explain the processes by which those experiencing long COVID attempt to alleviate cognitive difficulties and what they expect—or hope to receive—from the healthcare system. Ultimately, such insights may help inform more responsive rehabilitation services and guide future intervention development for this emerging patient population.

## 2 Methods

### 2.1 Study Design

We conducted a qualitative study grounded in Charmaz’s (2014) constructivist grounded theory methodology.^23^ This approach allows for inductive, iterative generation of theory from data, emphasising participants’ lived experiences and the social contexts shaping those experiences.^23^ We chose grounded theory because our aim was to not only describe patient experiences but also develop a conceptual understanding (a theory) of how individuals manage long COVID cognitive impairments and seek treatments. The study is reported in accordance with the 32-item COREQ (Consolidated Criteria for Reporting Qualitative Research) checklist, as shown in Appendix 1.

### 2.2 Participant Recruitment

Using purposive and theoretical sampling, we recruited 21 participants in the UK who either self-identified as having long COVID with prominent cognitive symptoms (such as “brain fog,” memory issues, or difficulty concentrating persisting for more than 12 weeks post-infection), or had this diagnosis confirmed by a physician. We specifically sought variation across age groups – targeting younger adults (aged 18–39 years) and older adults (aged ≥60 years) – to enable comparisons. These cut-offs were chosen because young adults under 40 typically represent early- to mid-adulthood, when work, education, and family responsibilities are most salient, whereas older adults aged 60 and above are more likely to be retired, have age-related comorbidities, and face a higher risk of age-associated cognitive decline.

Recruitment was conducted through the Long COVID Support charity (particularly via its COVID-19 Research Involvement Group) and the Lancaster University Centre for Ageing Research (C4AR) Panel. Individuals who expressed interest by contacting the principal investigator (D.S.) were screened to ensure they met the inclusion criteria: a prior confirmed COVID-19 infection, self-reported or physician-diagnosed cognitive difficulties related to long COVID (of any type) lasting at least three months, and the willingness and ability to discuss their experiences. Individuals who disclosed pre-existing cognitive impairments or a prior diagnosis of dementia were excluded to help ensure that reported difficulties were attributable to long COVID. We intentionally aimed for a roughly equal split between younger and older participants, anticipating that life stage might influence coping strategies and support needs. All participants provided informed consent online after receiving a participant information sheet and a study consent form. Ethical approval was obtained from the Lancaster University Research Ethics Committee (Ref. FHM-2025-4644-SA-2). Participants were assured of confidentiality and informed of their right to withdraw from the study at any point, up to two weeks after the interview.

### 2.3 Data Collection

We developed a semi-structured interview guide with open-ended questions, primarily informed by existing literature on long COVID.^17^ The draft guide was subsequently reviewed by two individuals with long COVID-associated cognitive impairments, who did not participate in the study but offered constructive suggestions for its refinement before the study commenced. Interviews were then conducted via secure online platforms, including Zoom and Microsoft Teams, with each session held as a one-on-one interview. Key prompts used during the interviews included: “Could you describe the cognitive problems you’ve had since long COVID (e.g., memory, concentration, etc.) and how they affect your life?”, “Were your long COVID-related cognitive issues diagnosed by a medical professional or are they self-perceived”, “What have you tried to help with these cognitive symptoms? – this could be strategies, treatments, anything – and how has that worked?”, “Have you sought medical help for these issues? What was that experience like?”, “What kind of treatment or support would you ideally want for the “brain fog” or memory troubles?”, and “Looking ahead, what are your hopes or expectations regarding your cognitive symptoms? Do you think they’ll improve, or is there something you are waiting for (like a specific therapy)?” We also asked targeted follow-ups based on age context (e.g. for younger participants: “How have cognitive issues affected your job or studies and what accommodations have you made or received?”; for older participants: “Did you worry at first that these problems were just ageing? How do family or friends view your cognitive symptoms?”). The guide remained flexible, allowing participants to raise additional relevant topics.

### 2.4 Data Analysis

Following Charmaz’s constructivist grounded theory approach, data collection and analysis were conducted iteratively.^23^ We began with initial line-by-line coding to capture participants’ in vivo expressions, followed by constant comparison across transcripts to identify similarities and variations. Focused coding was then used to cluster related codes into higher-level categories, which were subsequently refined through theoretical sampling and comparative analysis between age groups. Analytical memos supported the development of emerging ideas, and theoretical coding integrated these categories into a conceptual framework. The analytic process continued until theoretical saturation was reached in both age groups. More detailed information on the analytic procedures is available in Appendix 2. Rigour was ensured through two strategies: member checking, whereby a summary of the analysed results was sent to four randomly selected participants, who confirmed that the findings reflected their experiences; and triangulation with observations from patient support group discussions.

### 2.5 Patient and Public Involvement

Two individuals with lived experience of long COVID-associated cognitive impairment reviewed the draft interview guide prior to study commencement and provided constructive feedback to refine its content and wording. Four participants later took part in member checking, confirming that the preliminary analysis reflected their experiences. Although patients were not involved in recruitment or data collection beyond participation as study subjects, their contributions were central to shaping the interview process and interpretation of findings. The study results will be shared with participants and disseminated through long COVID patient support groups.

## 3 Results

### 3.1 Sample Characteristics

Among the 21 participants, 10 were aged 25–38 years (younger group; mean age: 32.5 years), and 11 were aged 60–72 years (older group; mean age: 65.2 years). The cohort comprised 14 women and 7 men. The mean interview duration was 48.4 minutes for younger participants (range: 41–62 minutes) and 51.7 minutes for older participants (range: 45–70 minutes). Table 1 summarizes the demographic characteristics of the study sample. Except for two individuals in the younger group whose symptoms were self-reported without formal medical confirmation, all other participants had physician-diagnosed long COVID-related cognitive impairment, established in either hospital or primary care settings. At the time of interview, cognitive symptoms had persisted for a mean duration exceeding 24 months (range: 15 months to 3.5 years). The mean duration was 19 months in the younger cohort and 30 months in the older cohort. Although many also reported concurrent long COVID manifestations—most commonly fatigue—cognitive dysfunction was consistently described as among the most disabling and persistent symptoms across these participants. In addition, none of the participants had a prior clinical diagnosis of dementia.

**Table 1.** Participant characteristics (N=21)

| **Table 1. Participant characteristics (N=21)** |  |  |
| --- | --- | --- |
| **Participants** | Young adults (n=10) | Older adults (n=11) |
|  | n (%) | n (%) |
| **Sex** |  |  |
| Man | 3 (30) | 4 (36.4) |
| Woman | 7 (70) | 7 (63.6) |
| **Age group (years)** |  |  |
| 18-39 | 10 (100) | / |
| ≥60 | / | 11 (100) |
| **Diagnostic methods for LCRCI** |  |  |
| Self-perceived | 2 (20) | 0 (0) |
| Physician-confirmed | 8 (80) | 11 (100) |

LCRCI, long COVID-related cognitive impairment

### 3.2 Theoretical Framework of Patient Adaptation to Cognitive Impairment

Using constructivist grounded theory analysis, we developed a theoretical framework that explains how individuals with long COVID cognitive impairments navigate their condition in the absence of definitive treatments (Figure 1). The core process can be described as “Striving for Agency in the Face of Cognitive Issues” – patients engage in proactive self-management and seek external support to regain a sense of control over their cognitive functioning, all while tempering their preferences and expectations based on the realities of long COVID and their personal life stage/context. Four major interrelated categories (themes) emerged, illustrated in Figure 1: “Adopting Self-Management Strategies (in Daily Life)”; “Healthcare Support (or Lack Thereof) and Desire for Validation”; “Treatment Preferences (Balancing Preferences for Non-Pharmacological vs. Pharmacological Interventions)”; and “Hope, Uncertainty, and Outlook (for the Future)”. Generational or age-related factors influenced each of these themes in nuanced ways, which are examined in detail in the following sections. Participant quotes are included to illustrate each theme in the following sections.

**Figure 1.**
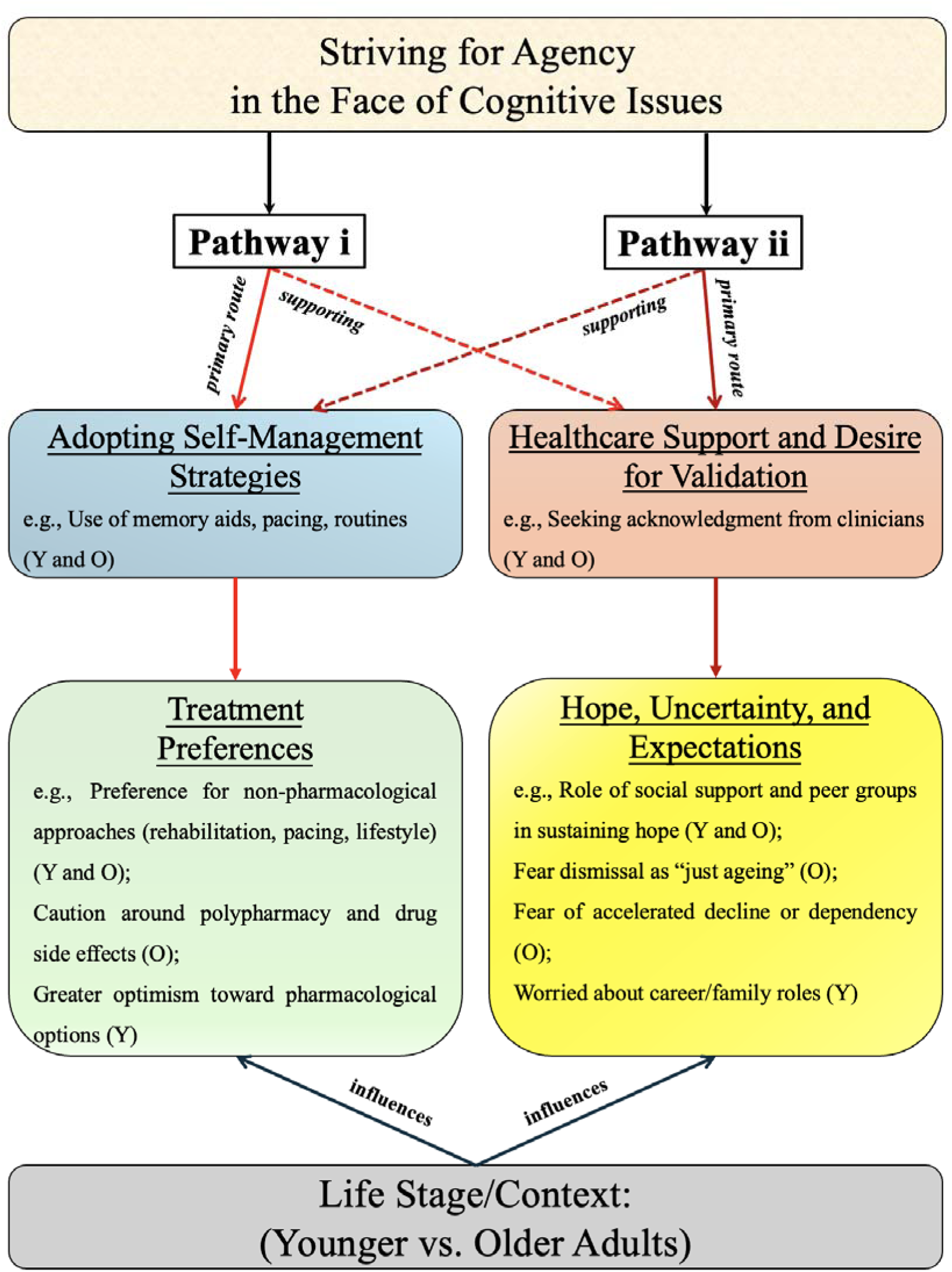
Constructivist grounded theory model of how participants with long COVID manage cognitive impairments through flexible pathways to agency.

The central process—striving to maintain or regain a sense of agency in the face of cognitive difficulties—is enacted through two interrelated and individually variable pathways. Some participants primarily engage in self-management strategies (e.g., pacing, memory aids, cognitive exercises), while others rely more heavily on navigating healthcare systems for support (e.g., medical consultations, referrals, or lack thereof). These two pathways may function as either a primary route or a supporting route depending on personal context, capacity, and resources. Together, these upstream processes influence treatment preferences (e.g., pharmacological vs. holistic approaches) and shape future-oriented expectations (e.g., hope for recovery vs. pragmatic adaptation). Life stage and contextual factors (e.g., younger vs. older adults) moderate how individuals prioritise, interpret, and act upon these processes. The model comprises four core categories (themes) with associated subcategories; Y denotes younger adults (18–39 years), and O denotes older adults (≥60 years).

#### 3.2.1 Adopting Self-Management Strategies in Daily Life

All participants described an array of practical strategies they had adopted to cope with their cognitive symptoms. Given the lack of proven drug therapies, they turned to behavioural and lifestyle adjustments to compensate for memory lapses, confusion, or slowed thinking. A strong sense of self-reliance was evident throughout their narratives. This theme encompasses how participants reorganised their routines, externalised information to memory aids, paced their activities, and exercised their minds, effectively creating personalised rehabilitation regimens.

Participants generally reported that these self-management efforts mitigated their cognitive difficulties to a degree, even though they did not eliminate them. Most participants reported that, over time, they had become more adept at managing their symptoms, which in some cases led to slight-to-moderate improvements in functional ability: “*At my worst, I was so forgetful I didn’t even feel safe working or driving*,” said an older adult (Man, 60s). He continued, “*I rely on loads of things now – reminders, alarms, my wife double-checking stuff – it keeps me going, just about*.” Younger participants often described this process as finding ways to adapt to or compensate for the cognitive difficulties associated with brain fog. While grateful for these workarounds, several also expressed frustration that they had to rely on them: “*I hate having to rely on a checklist all the time*,” one younger adult reflected, “*but at the moment, it’s the only thing that works*”(Woman, 30s).

Notably, there was no stark divide between younger and older adults in adopting most of these strategies – for example, most participants were using memory aids and pacing – but the triggers and contexts differed. Older adults, many of whom were retired, tended to focus their strategies on personal and household domains (remembering medications, avoiding cooking mishaps, managing bills with help). Younger adults discussed these as well but also emphasised strategies to maintain their work performance (like the software engineer’s extensive coding notes or the teacher’s half-days and checklist for class). In essence, younger individuals were often trying to hide or compensate for cognitive issues in professional settings, whereas older individuals were more concerned with maintaining independence in daily living. One older participant remarked, “*I’m lucky I’m not trying to hold down a job with this – I can’t imagine it. I just have to manage my home life, which is enough*” (Woman, 60s). In contrast, a younger participant said, “*I’m always setting up little routines at work, just so people don’t catch on that I’m struggling*” (Man, 30s). Another younger participant described practicing mindfulness meditation, which she felt helped calm her mind and improve her clarity of thought, illustrating how some participants explored psychosocial strategies alongside practical memory aids and routines. This underscores how life stage shaped the stakes: younger adults felt more external pressure to perform cognitively (e.g., career, parenting) and thus adopted sometimes elaborate coping systems to “pass” as fully functional, whereas older adults felt internal pressure to remain self-sufficient but had less obligation to keep complex schedules or perform multi-tasking beyond personal activities.

Nonetheless, the types of strategies overlapped considerably between age groups – lists, alarms, routines, avoidance of multitasking – suggesting that these are broadly useful cognitive adaptation techniques, aligned with what occupational therapists and clinicians recommend for managing post-COVID cognitive issues.^24^

#### 3.2.2 Healthcare Support and Desire for Validation

The second major theme centres on participants’ interactions with the healthcare system and how those experiences influenced their treatment journey. Many participants voiced some level of frustration or disappointment with the lack of concrete treatment options offered by healthcare providers. At the same time, many sought and appreciated validation – having a doctor acknowledge that their cognitive symptoms were real and part of long COVID (not just stress or ageing). This theme captures a dual sentiment: frustration at the lack of available treatments coexisted with appreciation that diagnostic confirmation and validation offered psychological reassurance. Participants also expressed yearning for more guidance and specialised care (e.g. cognitive rehabilitation programs), noting the current healthcare response felt insufficient relative to their needs.

In sum, participants expressed gratitude for validation and basic advice received, but clear dissatisfaction with the status quo of “no real treatment.” This fuelled a mix of self-reliance and advocacy: some had become vocal patient advocates calling for more research. There was a poignant contrast in some interviews between admiration for individual clinicians’ empathy and frustration at the system’s inertia. For instance, an older adult (Woman, 60s) said, *“My GP was very kind and actually knew about long COVID – that made a huge difference mentally. But neither he nor I are satisfied with ‘kindness’ as the end of it. We need effective treatments, and he agreed but said, we just have to wait for science. That’s hard to hear when you’re suffering now.”* This quote encapsulates the yearning in our sample: compassionate acknowledgement was appreciated but not enough – participants want tangible solutions from healthcare, and they want the healthcare system to catch up with their needs.

Notably, there were no major age differences in the frustration with lack of treatments; this sentiment was universal. However, older participants were more likely to mention concerns about doctors dismissing them (due to age or comorbidities), whereas younger participants more often mentioned needing validation in the workplace or academic settings. Both groups strongly desired more support: older adults, despite being mostly retired, wanted something to improve their cognition so they could retain independence and enjoy retirement, while younger adults wanted to get back to their prior productivity and fulfil roles (e.g., career, parenting). One slight difference was in healthcare-seeking behaviour: younger participants tended to push for referrals or research trials (some mentioned they had put themselves on waiting lists for long COVID research studies or consulted multiple doctors), whereas some older participants were less proactive in seeking care, at times resigning themselves to the perception that physicians had little more to offer after the initial consultation. For example, an older adult (Man, 60s) said, *“I haven’t bothered my GP again about it – I can tell they don’t have answers, and I just get on with it.”* In contrast, a younger adult (Woman, 30s) said, *“I kept going back and asking if there’s anything new we could try. I reckon they probably know me by my first name now.”* This might reflect generational attitudes towards medicine (older generations being more stoic or less expectant of a quick fix, younger generations being more proactive healthcare seekers). Nonetheless, all participants articulated a hope that healthcare would eventually offer more – as one younger adult put it, *“I’m hopeful that research will eventually come up with a treatment… I tell myself, maybe in a year or two they’ll figure out a drug or something. Waiting is hard, but at least it gives me some hope”* (Man, 20s).

#### 3.2.3 Balancing Treatment Preferences: Non-Pharmacological vs. Pharmacological Interventions

A prominent theme in our data was how participants perceived different modalities of treatment – essentially, what kinds of interventions they preferred or were willing to try, and which they were sceptical about. Since no standard treatment exists, participants were left to weigh self-directed lifestyle measures vs. medications or other biomedical interventions (should any become available or accessible). We found that these participants continuously balanced these options, often favouring a conservative, wellness-focused approach by necessity, but also expressing openness or hope towards medical therapies if proven safe and effective.

In terms of age differences, as noted, older adults generally prioritised non-drug approaches and had more scepticism about pharmacological solutions, whereas younger adults were somewhat more optimistic about pharmacological treatments (willing to consider off-label prescriptions, clinical trials, etc.). Older participants frequently cited concerns like polypharmacy (many were already on medications for hypertension, hyperlipidaemia, etc., and did not want to add risky new ones). Younger participants voiced more concern about time – they did not want to wait years for relief – making them more inclined to pursue something if available sooner. Despite this, both groups ultimately said they would choose whatever approach gave them their cognitive function back. *“I’ve nothing against medication – if a trial came out tomorrow showing an anti-inflammatory made a real difference with brain fog, I’d be up for trying it,”* said a younger adult (Man, 20s). *“But as nothing’s been proven yet, I’m sticking with diet, exercise and the like.”* Older adults said very similar things with roles reversed: *“I’m focusing on lifestyle and pacing, but if they do find a medication that truly helps and it’s safe, I’ll happily take it. I just don’t buy that there’s a magic bullet yet,”* (Woman, 60s). In practice, most participants were already engaging in non-pharmacological strategies and were waiting for pharmacological options to emerge.

An interesting subtheme was participants’ attitudes towards specific alternative therapies or supplements. A few mentioned things like antihistamines, omega-3, or herbal supplements that circulate in patient forums as potential supports. Generally, some participants tried vitamins and supplements (many took vitamin D, B, omega-3 as noted), but they were sceptical of unproven “miracle cures” sold online. *“There’s a lot of quackery out there claiming to cure long COVID brain fog,”* one older participant warned (Woman, 60s). *“I saw something about hyperbaric oxygen – I’d probably give it a go if I could afford it, even though it’s experimental. But expensive supplements without evidence? No chance, I’m not wasting my money.”* This indicates a discerning approach; indeed, a few participants explicitly criticised profiteering and misinformation. One participant (Woman, 60s) said, *“I’ve been approached by people trying to sell nootropic supplements. I’m not falling for that nonsense. Let’s see some actual research first.”* So while desperate for solutions, participants navigated a careful line, preferring to conserve trust for credible avenues (i.e., medical trials, NHS clinics) rather than fringe remedies. Younger participants in our sample were somewhat more likely to mention hyperbaric oxygen therapy (HBOT) as something they would consider if accessible: *“I’d consider HBOT – I’m open to trying things, as long as they’re safe,”* said a younger adult (Man, 30s). Indeed, he and others had read news of small studies suggesting HBOT might improve cognitive symptoms.^25^ However, cost could be a barrier (given that HBOT is not provided in public healthcare for long COVID, and private sessions are often expensive). Overall, participants prioritised low-cost, low-risk measures first (i.e., diet, pacing, exercise within limits, and mental exercises) and treated potential high-cost or higher-risk interventions as a distant secondary option to be approached carefully or ideally through research channels.

#### 3.2.4 Hope, Uncertainty, and Outlook for the Future

This theme captures participants’ reflections on the future course of their cognitive impairments – their hopes, fears, and overall outlook. It became evident that managing long COVID-related cognitive impairment is not just a practical endeavour but also a psychological one: patients grapple with uncertainty about whether they will ever fully recover and what that means for their identities and lives. This theme includes the emotional coping dimension and how age and life stage influence one’s future orientation.

In terms of age influences here, younger adults tended to emphasise the desire to return to their pre-illness selves, often struggling to envisage accepting a new baseline, whereas older adults more frequently described the need to reconfigure their expectations of ageing in light of these symptoms. A few older participants explicitly voiced fears that their long COVID cognitive problems might signal the onset of dementia, worrying that “brain fog” could represent accelerated cognitive ageing. Yet both groups exhibited resilience: most participants in our sample had found some way to keep going, whether through faith that improvement will come or through reframing life to accommodate limitations. Strength of social support often determined outlook: those with understanding families or support groups were generally more hopeful, as they felt less alone and had help in daily life. Conversely, a couple of participants who lived alone or felt misunderstood admitted to dark periods of hopelessness. *“There were days I thought, is this really worth it – I felt like such a mess, unable to string a thought together,”* one younger adult confided (Woman, 30s), alluding to depressive thoughts. She then added, *“Talking to others changed that. Now I have hope again – maybe not a full recovery, but I can still find joy and purpose even if my brain isn’t what it was.”* This transformation underscores how peer and community support may help convert despair into a more hopeful acceptance for many.

In conclusion, participants’ expectations for the future were a dynamic interplay of hope and realism. No one had completely given up, though some older participants came close after repeated setbacks. Likewise, no one was blindly optimistic that a cure was around the corner – they were aware recovery might be slow and incomplete. Instead, most cultivated a mindset of cautious optimism—maintaining hope for medical advances or gradual natural improvement, while simultaneously preparing to adapt to the condition in order to preserve their current level of functioning and prevent further decline. This mindset was useful practically, as it motivated them to continue self-management and advocacy (agency) without succumbing to despair. In the grounded theory model, this theme of managing hopes, uncertainty, and expectations is what ties together their ongoing efforts (other core themes) with their emotional well-being. Despite differences in life situations, younger and older participants shared a fundamental resilience: a determination to either improve or adapt, and in most cases, both. As one participant wisely put it, *“I tell myself, ‘Maybe tomorrow will be a bit better.’ Even if it isn’t, a hopeful mindset gets me through the day”* (Man, 60s).

## 4 Discussion

### 4.1 Navigating Cognitive Challenges in Long COVID Across Age Groups

In this constructivist grounded theory study, we developed a comprehensive understanding of how UK adults – both young and older – navigate the challenge of cognitive impairments in long COVID. Our findings highlight several key insights. First, participants have overwhelmingly turned to self-management strategies to cope with long COVID-related cognitive impairments (often described as “brain fog”), establishing rigorous routines, using memory aids, pacing their mental activities, and engaging in cognitive exercises. These patient-driven adaptations align with emerging clinical recommendations that emphasise energy conservation and compensatory techniques for long COVID.^17^ The extent of self-management reported in our study underscores that, in the absence of formal treatments, participants essentially improvise their own cognitive rehabilitation programs at home. This resonates with previous qualitative observations that long COVID patients resort to trial-and-error self-care and peer advice due to unmet needs in the healthcare system.^21^ Our study extends those observations by detailing the specific tactics used for cognitive symptoms and by comparing age groups. We found that while younger and older adults adopted similar tools – such as lists, alarms, structured routines – these were applied in different life contexts (workplace vs. home management), and each age group faced unique challenges in implementation (e.g. younger individuals hiding their aids at work to avoid stigma, older individuals learning to use new technologies for reminders). Notably, many participants in both groups reported meaningful functional improvements through these compensatory strategies, even if their underlying cognitive impairment persisted. This suggests that widespread dissemination of occupational therapy techniques and cognitive compensation skills may benefit the broader long COVID population.^17^ Health services should consider offering training on such strategies – for instance, memory skills workshops or fatigue management programs – which many participants in our study found helpful when available. Encouragingly, some participants independently developed techniques akin to those recommended for mild cognitive impairment or traumatic brain injury rehabilitation (e.g. external memory aids, avoiding multitasking).^17^ Future research might formalise these patient innovations into structured cognitive rehabilitation interventions for long COVID, which could then be tested for efficacy.

Second, our results illuminate the profound frustration with the current lack of medical treatments, but also the crucial importance of validation and support from healthcare providers. Participants did not expect their GP to magically cure long COVID “brain fog” – they understood the novelty of the condition – but they deeply valued being heard and not dismissed. Sadly, some older participants in our study initially encountered ageist or sceptical attitudes (e.g. symptoms attributed to ageing or anxiety), reflecting what other reports have noted as a tendency to minimise or misattribute long COVID patients (especially older adults) in the early pandemic period.^26,27^ However, many eventually found clinicians who acknowledged their cognitive issues as part of long COVID, which often provided psychological relief by validating that their experiences were genuine and shared. This validation is more than a courtesy; prior qualitative work on long COVID emphasises that believing patients and legitimising their illness experience can improve their mental health and engagement with care.^17^ Our study reinforces that: participants who felt validated described less self-blame and despair. Healthcare professionals should receive training to actively validate and listen to long COVID patients – a seemingly small intervention that may mitigate feelings of isolation and emotional turmoil that many described before they received a formal diagnosis. Alongside validation, participants desired more tangible support: most participants expressed a need for structured cognitive rehabilitation services and/or guidance on treatment options. The majority of our sample had only been told to pace themselves and were left to find solutions alone, which parallels findings that long COVID patients often feel abandoned by healthcare and forced into self-care.^28^ The implication for practice is that health systems should move beyond mere recognition to providing resources. For example, integrating neuropsychologists or occupational therapists into long COVID clinics could allow for formal cognitive evaluations and tailored strategy training – an approach that participants in our study explicitly desired, drawing parallels to the cognitive rehabilitation routinely offered to stroke patients. Additionally, given the lack of approved medications, clinicians should be prepared to discuss off-label or experimental options openly and empathetically with patients. We found that some younger, well-informed participants were exploring off-label treatments (e.g. stimulant or anti-inflammatory medications) on their own. This highlights a need for clear, evidence-based guidance that clinicians can share about which interventions are safe to try and which are not recommended outside trials. In our study, those who did try experimental medications (e.g., Low Dose Naltrexone, LDN) did so with medical supervision after their own research. Not all patients will have the resources or confidence to broach such topics – thus, clinicians might take the initiative to stay updated on emerging trials and be willing to have nuanced conversations about risk vs. benefit of off-label therapies, rather than a flat dismissal.

Third, our study sheds light on treatment preferences among long COVID patients – an area previously unexplored. We found a general leaning towards non-pharmacological interventions in principle, yet an openness to medication if it proved effective. Essentially, patients conveyed that while they were willing to adopt lifestyle-based strategies, they would also welcome the availability of effective pharmacological treatments to improve their cognitive function. This balanced view is important for researchers and policymakers to note. On one hand, it underlines that investments in non-drug approaches (like rehabilitation programs, mindfulness training, fatigue management classes) will be embraced by patients and should be expanded. Indeed, many participants in our study who accessed such services reported benefits (e.g. the patient who learned mindfulness to calm her mind and felt it helped her think clearer – consistent with prior research suggesting meditation may improve attention).^29^ On the other hand, participants clearly hold hope for biomedical solutions and many are willing to volunteer in trials – a finding that aligns with surveys where long COVID patients report high interest in experimental therapies and research participation.^30^ Notably, older adults in our sample were more cautious about pharmacological treatments than younger adults, citing fear of potential side effects and a general preference for “natural” coping. This suggests that age-tailored communication about treatments is needed: older patients may need extra reassurance and information on safety and may prefer starting with lower-intensity interventions. Younger patients, conversely, might be more willing to try aggressive therapies to regain high-level function, but may require guidance to channel that eagerness safely (for example, enrolling in clinical trials rather than self-medicating with unproven drugs). Notably, both age groups strongly agreed that any treatment must be evidence-based (or at least plausibly beneficial) – they were wary of “snake oil” and internet hype. This underscores the importance of combatting misinformation in the long COVID space and accelerating rigorous research. Encouragingly, large-scale trials are now underway (e.g. the UK’s STIMULATE-ICP trial and US NIH’s RECOVER trial) to test various interventions from antihistamines to metformin.^31,32^ Our study participants’ perspectives support such efforts – patients would like to see scientific progress and are eager to know the results, so that their care can shift from purely supportive to targeted. In practice, as trial evidence emerges, clinicians should be prepared to update patients and implement new treatments relatively quickly, because the pent-up demand is potentially enormous. This is one advantage of maintaining an ongoing relationship and trust: patients who feel their providers are on their side will look to them for latest treatment options rather than seeking risky unverified remedies elsewhere.

Fourth, our study reveals how coping with cognitive impairment in long COVID is not only a matter of managing memory or concentration – it is intimately tied to identity, mental health, and social roles. Participants across ages described a sense of loss: loss of the “sharp” person they used to be, loss of independence or professional identity, and in some cases loss of confidence and self-worth. Similar experiences are reflected in other qualitative studies–for example, a study noted that “brain fog” could lead to profound identity disruption and anxiety in long COVID sufferers, as they could no longer trust their minds or perform normal tasks.^20^ We extend this knowledge by showing that such identity struggles manifest somewhat differently by age: younger adults in our sample felt “robbed” of their prime years, often grieving lost career momentum or parenting capacity, whereas older adults grieved a more sudden end to their career or active retirement, or feared accelerated entry into dependency, with some explicitly worrying that their cognitive problems might accelerate dementia. Both required some degree of psychosocial adjustment. Many participants described reaching a level of acceptance – reframing priorities, adjusting self-expectations, and focusing on what they can still do. This reflects a coping trajectory often seen in chronic illness adaptation, moving from resistance to integration of the illness into one’s life story.^17^ Clinicians, especially in primary care and mental health, should support patients in this adaptation process. This might include referral to counselling or support groups. In our study, those who engaged with peer support reported it to be profoundly helpful – a finding consistent with numerous reports that peer communities (online or in-person) have been a lifeline for long COVID patients.^33^ Healthcare providers can encourage patients to connect with reputable support groups (while cautioning against groups that may spread misinformation). Additionally, integrating mental health professionals into long COVID care teams could help address the depressive and anxious feelings that accompany cognitive dysfunction.^34^ Several participants in our cohort mentioned benefiting from therapy or mindfulness in coping with the emotional aspect. Notably, one theme that arose was participants deriving hope from feeling they are contributing to solutions – for instance, taking part in our research or helping others in forums. This aligns with the concept of post-traumatic growth,^35^ or finding meaning through adversity. Clinicians might leverage this by, for example, inviting patients to share successful tips with others (as some clinics do by hosting group sessions) or involving patient representatives in co-designing long COVID services. Doing so can reinforce patients’ sense of agency and value, counteracting the disempowerment many described when their brain would not cooperate.

Additionally, while many participants in our study demonstrated resilience, wider community reports highlight that despair might escalate into severe anxiety and depression, as well as, in some cases, suicidal ideation.^36^ This risk may be particularly acute when patients feel unsupported or dismissed by healthcare professionals. Hence, recognising and addressing these mental health risks should be a core component of long COVID care, underscoring the need for early psychological assessment and access to crisis support.

### 4.2 Implications for research

Our study highlights several research gaps. One is the urgent need for clinical trials of interventions targeting cognitive impairment in long COVID. Participants in our sample were essentially acting as “citizen scientists,” experimenting with supplements or off-label drugs and collectively noting what helps. Systematic research could validate or refute these anecdotal remedies – for example, trials of stimulant medications (e.g., methylphenidate, modafinil) for long COVID cognitive function are still lacking and warranted, given their use supported by emerging evidence in analogous conditions like chemo-brain or traumatic brain injury.^37^ Another gap is studying cognitive rehabilitation protocols formally in long COVID. While such programs exist for other neurological conditions, they should be tailored and tested for post-COVID patients – perhaps delivered via telehealth given many patients’ mobility limits.^38^ Our participants expressed enthusiasm for participating in such rehabilitation programs if offered, indicating promising prospects for recruitment into intervention studies. Additionally, research should explore the long-term trajectory of cognitive impairments: many in our older group wondered if long COVID “brain fog” accelerates cognitive ageing or dementia risk. Epidemiological studies with follow-up neuropsychological testing could address this, thereby informing the level of monitoring and preventive care needed for older COVID survivors.^39,40^ At the same time, patients with long COVID-related cognitive impairment may also express concern that without timely intervention or monitoring, including the use of neuroimaging, a critical window for preventing irreversible cognitive damage could be missed. This highlights the importance of investigating not only long-term trajectories but also early markers and interventions that could mitigate permanent harm. Finally, further qualitative research could examine other demographic influences, such as gender (e.g., do women and men cope differently with “brain fog”?) or socioeconomic status (e.g., patients with fewer resources might face different challenges, such as having less flexible jobs or difficulty accessing care). Another potentially important factor is cognitive reserve: individuals with higher educational attainment or more intellectually and socially engaged lifestyles may possess greater resilience against cognitive decline. Given that many of our participants reported using compensatory strategies such as pacing or memory aids, future studies could investigate how differences in cognitive reserve influence long COVID trajectories and whether interventions to enhance reserve (e.g., cognitive training, social engagement) might mitigate the long-term impact of “brain fog.”^41^

### 4.3 Implications for clinical practice

Based on our findings, we offer the following practice recommendations:

1. Acknowledge and validate: when patients report cognitive issues, clinicians should explicitly affirm that these are real and common in long COVID and not a personal failing. Even brief reassurance that such symptoms are a recognised component of long COVID, and not merely psychological in origin, may help to alleviate patient distress.
2. Provide practical support: refer patients to occupational therapy for cognitive strategy coaching if available; if not, clinicians can share basic tips (e.g., use of planners, setting alarms, breaking tasks into smaller steps), which our participants found helpful. Clinicians may also help by discussing low-risk options with potential benefit and supervising their use, reducing the sense that patients must experiment entirely on their own.
3. Multidisciplinary care: ensure long COVID services include psychology/psychiatry input to help patients with adjustment, especially those struggling with identity loss or anxiety about cognitive decline. Techniques from brain injury rehabilitation and chronic illness therapy (e.g. cognitive restructuring, stress management) may be adapted here.
4. Keep patients informed: given their appetite for knowledge, clinicians should stay up-to-date on long COVID research and share new findings or trial opportunities with patients (for example, mentioning when a relevant trial opens or results come out, rather than waiting for patients to ask). This partnership approach will enhance trust and align care with patient expectations.
5. Individualise by age: for younger patients, ask about work and provide documentation or occupational health engagement to facilitate accommodations (our younger participants benefited when employers were informed and flexible). Recognise, however, that employment and occupational roles can be important across all ages. For older patients, it may be important to screen for underlying mental health concerns as well as potential early cognitive decline. Formal cognitive testing can be useful for both younger and older patients, either to provide reassurance or to identify emerging difficulties.

### 4.4 Strengths and Limitations

To our knowledge, this is the first qualitative study specifically examining treatment preferences and expectations for cognitive impairments in long COVID in the UK, and the compare experiences and expectations across age groups. A strength of our study is the inclusion of both older and younger adults, which allowed us to identify age-related nuances that single-cohort studies might miss. We used a rigorous grounded theory methodology, with constant comparison and iterative sampling until reaching theoretical saturation, enhancing the credibility of our findings. The use of rich interview data enabled us to construct a grounded theory that is well-anchored in participants’ own perspectives. We also triangulated our findings with observational insights from patient support group discussions, in addition to participant validation (member checking by four interviewees), to improve trustworthiness.

However, limitations must be acknowledged. Our sample, while diverse in age and gender, was relatively homogenous in other aspects – all were from the UK and the majority were White British. Cultural and ethnic factors might influence how cognitive symptoms are perceived and managed (for instance, stigma around cognitive issues or different health-seeking behaviours), so our findings may not fully transfer to long COVID populations in other cultural contexts. Further research including a more ethnically and internationally diverse sample would be valuable. Additionally, our study relied either on participants’ self-perceived cognitive impairments or on clinical diagnoses confirmed by hospitals or primary care physicians; we did not conduct objective neurocognitive tests during the interviews. It is possible that what patients labelled “brain fog” encompasses a range of specific deficits (attention vs. memory vs. executive function),^12^ which might have different impacts and coping strategies. Future studies could integrate qualitative interviews with cognitive assessments to correlate subjective experiences with objective profiles.

Another limitation is potential recall bias or self-selection bias: those who volunteered to be interviewed might be more proactive and oriented towards coping (indeed, many of our participants had actively implemented strategies and sought information). Long COVID patients who are more isolated or who have not found effective coping mechanisms might voice even greater frustration or hopelessness, which our theory may underrepresent. Finally, given the rapidly evolving nature of long COVID research, participants’ expectations might change as new knowledge emerges – our data only capture a specific moment in the post-pandemic era. Ongoing qualitative research is needed to see how patient perspectives shift with developments such as clinical trial results or improved service provision.

## 5 Conclusion

This grounded theory study identifies “Striving for Agency in the Face of Cognitive Issues” as the central process through which people with long COVID-related cognitive impairment manage their condition. In the absence of proven treatments, participants worked to preserve autonomy and identity by adopting self-management strategies, seeking validation, and negotiating uncertain healthcare encounters. Younger adults expressed this striving through proactive pursuit of clinical trials and experimental treatments, while older adults emphasised pragmatic adaptation and maintenance of independence. For primary care and wider health systems, recognising and supporting this striving for agency is critical. Services must rapidly evolve towards an integrated, holistic, and patient-centred model structured around three essential pillars: validation (listening to and legitimising patient experiences), rehabilitation (delivering accessible and evidence-based cognitive support), and innovation (accelerating research and translation of new therapies). This comprehensive framework must directly address specific patient-identified needs, including specialized memory clinics, tailored cognitive rehabilitation programs, workplace accommodations, peer support networks, and compassionate, individualized care. Ultimately, respecting and enhancing patient agency, promptly addressing patient-expressed needs, and rapidly translating research advancements into clinical practice can immediately alleviate patient suffering, empower individuals experiencing cognitive impairment, and significantly advance collective progress in confronting the emerging frontier of post-viral illness.

## Acknowledgements

We sincerely appreciate all study participants for their time and valuable insights, which significantly enriched the quality and depth of this research. We also thank the two individuals with long COVID-associated cognitive impairment who reviewed our initial interview guide and offered constructive suggestions prior to study commencement, as well as the four participants who took part in member checking to confirm that our analysed results reflected their experiences.

## Authors’ contributions

DS: Conceptualization, Methodology, Investigation, Data collection and curation, Formal analysis, Visualization, Project administration, Writing-original draft preparation, and Writing-review and editing. TC and CH: Conceptualization, Methodology, Writing-review and editing, and Supervision. All authors read and approved the final manuscript.

## Data availability

The data are not publicly available as they contain information that could compromise the anonymity of research participants.

## Ethics approval

Our study strictly adhered to the ethical guidelines outlined in the Declaration of Helsinki (2013 revision) throughout all stages of the research, with necessary approvals obtained from Lancaster University’s Faculty of Health and Medicine Research Ethics Committee (FHM-2025-4644-SA-2). All participants provided written informed consent before participating in the interviews.

