## Appendix 2 for "Patient needs and expectations regarding cognitive impairments in long COVID: perspectives of young and older adults in the UK"

**This appendix provides detailed information on the grounded theory coding and analysis procedures employed in the study *‘Patient needs and expectations regarding cognitive impairments in long COVID: perspectives of young and older adults in the UK.’* It outlines the iterative coding process, category development, and theoretical modelling approach, following Charmaz’s (2014) constructivist grounded theory methodology.**

Following Charmaz’s (2014) constructivist grounded theory approach, data collection and analysis proceeded iteratively. Transcripts were imported into *NVivo* for organization. We began with initial coding – a line-by-line examination of each transcript, assigning provisional codes to capture in vivo terms and actions (e.g. “mind goes blank mid-task”, “uses phone alarms”, “doctor ruled out dementia”, “feels like a burden”). As coding progressed, we used constant comparison – comparing codes to other codes and incidents within and across transcripts – to identify patterns and variations. We wrote analytical memos to explore emerging ideas (for instance, noting that several participants described rigid routines as coping mechanisms, and comparing how younger vs. older individuals spoke about routines).

Next, we engaged in focused coding, wherein we grouped the numerous initial codes into higher-level categories based on conceptual similarity and significance to the research questions. For example, codes like “writing everything down,” “using to-do lists,” and “externalizing memory” were clustered under a category we labelled “Compensatory memory aids”, while codes relating to interactions with healthcare (e.g. “GP didn’t have answers,” “attended long COVID clinic,” “felt validated by diagnosis”) formed a category around “Healthcare response and support”. We constantly compared data within each category to ensure it coherently represented participants’ experiences and to check for negative cases or exceptions. Through this process, several key categories/themes solidified: “Adopting Self-Management Strategies” (encompassing cognitive and lifestyle adaptations), “Healthcare Support and Desire for Validation” (including both positive validation and frustration/lack of solutions), “Treatment Preferences” (participants’ inclinations toward or against certain treatment modalities, e.g. medication vs holistic therapy), and “Hope, Uncertainty, and Outlook.” We also explicitly compared younger and older participants’ data at this stage, looking for age-related contrasts within each category. This led to memos on generational perspectives (for instance, noticing older adults often emphasized acceptance and caution with experimental treatments, whereas younger adults expressed urgency to regain cognitive normalcy). We continued interviewing new participants strategically to fill out these categories and test our emerging understandings (theoretical sampling). For instance, after noticing a potential age difference in attitudes to medication, we sought additional interviews in each age group to probe that issue further.

Finally, we proceeded to theoretical coding and model construction. We examined how the major categories related to each other and to the core phenomenon (managing cognitive impairment in long COVID). A tentative central storyline emerged: patients seeking to reclaim cognitive function through self-driven means in an environment of medical uncertainty, moderated by personal context (age/life stage). We integrated the categories into a conceptual framework, wherein “Adopting Self-Management Strategies” and “Healthcare Support (or lack thereof) and Desire for Validation” feed into an ongoing process of striving for control or improvement, all occurring under the umbrella of limited treatment options and the need to adjust expectations. We identified “Striving for Agency in the Face of Cognitive Issues” as a unifying concept – patients actively try to help themselves and maintain hope, even as they come to terms with possible long-term changes. We achieved theoretical saturation within each age group when additional interviews yielded no fundamentally new codes or relationships – by the 10th interview in the younger group and the 11th in the older group, participants were echoing previously observed concepts, with only nuanced additions.

To enhance the methodological rigor of the analysis, we implemented several validation strategies, including member checking—whereby synthesized findings (including the final theoretical framework) were shared with four randomly selected participants to ascertain the extent to which the interpretations resonated with their lived experiences—and triangulation, achieved through cross-referencing interview data with observational insights drawn from patient support group discussions.
